# Revisiting IgG antibody reactivity to Epstein-Barr virus in Myalgic Encephalomyelitis/Chronic Fatigue Syndrome and its potential application to disease diagnosis

**DOI:** 10.1101/2022.04.20.22273990

**Authors:** Nuno Sepúlveda, João Malato, Franziska Sotzny, Anna D Grabowska, André Fonseca, Clara Cordeiro, Luís Graça, Przemyslaw Biecek, Uta Behrends, Josef Mautner, Francisco Westermeier, Eliana M Lacerda, Carmen Scheibenbogen

## Abstract

Infections by the Epstein-Barr virus (EBV) are often at the disease onset of patients suffering from Myalgic Encephalomyelitis/Chronic Fatigue Syndrome (ME/CFS). However, serological analyses of these infections remain inconclusive when comparing patients with healthy controls. In particular, it is unclear if certain EBV-derived antigens eliciting antibody responses have a biomarker potential for disease diagnosis. With this purpose, we re-analysed a previously published microarray data on the IgG antibody responses against 3,054 EBV-related antigens in 92 patients with ME/CFS and 50 HCs. This re-analysis consisted of constructing different regression models for binary outcomes with the ability to classify patients and HCs. In these models, we tested for a possible interaction of different antibodies with age and gender. When analyzing the whole data set, there were no antibody responses that could be used to distinguish patients from healthy controls. A similar finding was obtained when comparing patients with noninfectious or unknown disease trigger to healthy controls. However, when data analysis was restricted to the comparison between HCs and patients with a putative infection at disease onset, we could identify stronger antibody responses against two candidate antigens (EBNA4_0529 and EBNA6_0070). Using antibody responses to these two antigens together with age and gender, the final classification model had an estimated sensitivity and specificity of 0.833 and 0.720, respectively. This reliable case-control discrimination suggested the use of the antibody levels related to these candidate viral epitopes as biomarkers for disease diagnosis in this subgroup of patients. When a bioinformatic analysis was performed on these epitopes, it revealed a potential molecular mimicry with several human proteins. To confirm these promising findings, a follow-up study will be conducted in a separate cohort of patients.

## Introduction

Infections by the ubiquitous Epstein-Barr virus (EBV) are linked to multiple sclerosis, rheumatoid arthritis, systemic erythematosus lupus, lymphomas, among other known diseases (1–3). A less-known disease where EBV infections are particularly relevant is Myalgic Encephalomyelitis/Chronic Fatigue Syndrome (ME/CFS) (4–6). The hallmark symptom of this condition is an unexplained but persistent fatigue that cannot be alleviated by rest and that can be exacerbated by minimal physical and emotional effort (7,8). In ME/CFS, acute EBV infections are reported by a subset of patients at the onset of their symptoms (9,10). Reactivation of latent EBV infections has also been described during the disease course (11). However, the existing evidence remains inconclusive on whether the prevalence of these reactivations is either higher or lower in patients than in healthy controls (5). This conflicting evidence notwithstanding, ME/CFS patients show deficient B- and T-cell responses against EBV and altered antibody profiles when compared to healthy controls (10,13–15). Finally, CD4+ T cells recognizing self-peptides on HLA-DR15, the strongest genetic risk factor for multiple sclerosis, have been shown to cross-react with peptides derived from EBV (16). Multiple sclerosis patients share many symptoms with the ones suffering from ME/CFS (17– 19). EBV antigens were also reported to share sequence homology with human peptides derived from the myelin basic protein (20–22), lactoperoxidase (23), and anoctamin-2 (24,25). These observations suggest that molecular mimicry between human and EBV-derived antigens could play a role in the pathogenesis of ME/CFS. This is in line with the recent hypothesis that has linked ME/CFS pathogenesis with chronically activated immune responses (26). Our assumption raises the possibility that the immune system of some ME/CFS patients is oscillating between an activation state that attempts controlling latent herpesviruses infections and the suppression of deleterious autoimmune responses via activation of regulatory T cells (26). Thus, considering the growing body of evidence that links EBV infection with the pathogenesis of ME/CFS, studies that aim at elucidating underlying mechanisms are certainly needed.

A major problem in investigating ME/CFS is the inexistence of a robust biomarker that could ascertain the disease diagnosis. In the past, different discovery studies suggested certain cytokines, antibodies against self and non-self antigens, microRNAs, and methylation markers as potential disease biomarkers (27). Antibodies against EBV antigens are of particular interest as disease biomarkers given the above described evidence connecting this virus with the disease and routine application of serological assays in the clinical practice. However, EBV antigens included in commercial kits are mostly markers of exposure to the infection and are unable to distinguish between patients with ME/CFS and healthy controls (28). This distinction can only be made when comparing a subset of clinically diagnosed ME/CFS patients with an EBV infection trigger to healthy controls (10). A serological evaluation of antibodies against less-studied EBV antigens did not identify any that could be used as a specific disease biomarker (29). However, this antibody evaluation was done using a limited number of EBV-derived antigens and no subgroup analysis was performed. The lack of patients’ stratification in ME/CFS studies reduces the chance of reproducing the same findings in follow-up studies (27,30). Therefore, it is still possible to identify alternative antigens whose antibody responses could be used as disease biomarkers for a subgroup of patients.

Recently, we analyzed antibody responses against about 3,000 overlapping antigens derived from 14 EBV proteins (23). The aim of this original study was to extract an antibody signature against EBV in ME/CFS patients when compared to healthy controls. In the present study, we extended the analysis of the obtained data with the specific objective of optimizing biomarker discovery. In particular, we compared patients with or without an infectious trigger at disease onset to healthy controls in order to discover EBV-derived antigens whose antibody responses could be used for ME/CFS diagnosis.

## Materials & Methods

### Study participants

Ninety-two ME/CFS patients were recruited between 2011 and 2015 at the Charité outpatient clinic for immunodeficiencies at the Institute of Medical Immunology in the Charité Universitatsmedizin Berlin, Germany. Additional fifty individuals were recruited from the employees of the same clinic, who self-reported to be healthy and to not suffer from fatigue. However, neither clinical nor laboratory assessment was performed to confirm the healthy status of those individuals. ME/CFS patients and healthy controls were matched for gender and age (Table 1) with 50% of women and an overall average age of ∼43 years of age. Fifty-four out of 92 patients (58.7%) reported an acute infection at their disease onset, whilst the remaining 38 patients (41.3%) reported either a disease trigger other than infection, did not know their disease onset or the information about the disease trigger was missing. These two subgroups were also matched for age and gender (Table 1).

**Table 1.**
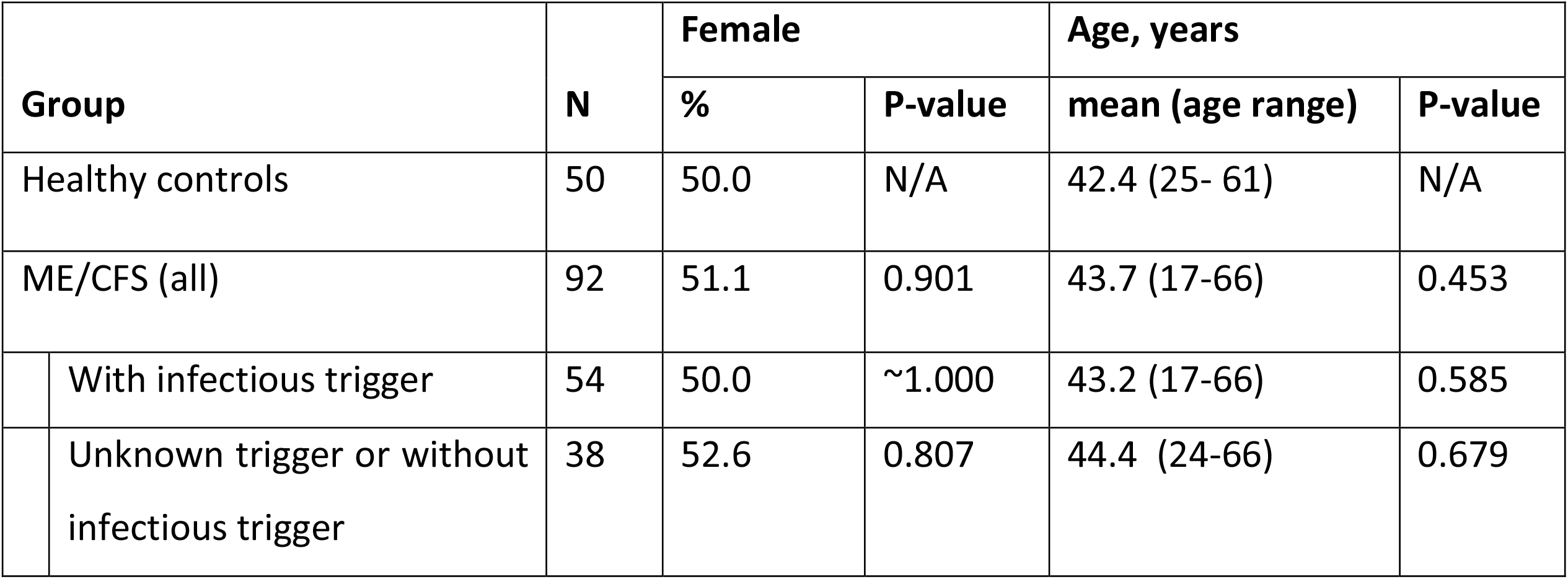
Basic characteristics of ME/CFS patients and healthy controls and their statistical comparison, where p-values refer to the comparison between ME/CFS groups and healthy controls

### Peptide Array

Data under analyses refer to signal intensities derived from IgG antibody responses to 3,054 EBV-associated peptides measured by a seroarray described in detail in the original study (23). These peptides consisted of partially overlapping 15 amino acids and covered the full length of the following proteins (Supplementary Table 1): BALF-2, BALF-5, BFRF-3, BLLF-1, BLLF-3, BLRF-2, BMRF-1, BZLF-1, EBNA-1, EBNA-3, EBNA-4, EBNA-6, LMP-1, and LMP-2. The peptides covering these antigens were 15 amino acids (15-mer) in length and overlapped in 11 amino acids. The amino-acid sequences of these peptides were representative of the following EBV strains: AG876 (West Africa, EBV type 2), B95-8 (USA, EBV type 1), GD1 (China, EBV type 1), Cao (China, EBV type 1), Raji (Nigeria, EBV type 1), and P3HR-1 (Nigeria, EBV type 2). These data are publicly available as a supplementary table of the original study (23).

### Statistical analysis

We used the Chi-square test to compare the gender distribution between ME/CFS patients and healthy controls. The non-parametric Mann-Whitney test was used to compare the medians of the respective age distributions. There was evidence for age-and gender-matched distributions if the p-values of these tests were greater than the significance level of 0.05.

We first performed a multivariate analysis using (i) the classical principal component analysis (PCA) and (ii) computing different correlation matrices using Spearman’s correlation coefficient (which is invariant to monotonic changes in the scale of the data and is robust against the presence of outliers, and does not depend on the normality assumption). We then performed linear discriminant analyses (LDA) to determine the best linear combination of all antibody responses that could distinguish ME/CFS patients and their subgroups from healthy individuals. A similar analysis was done to compare the two subgroups of ME/CFS patients. The outcome of each LDA was the estimated classification probability for every individual. These classification probabilities were then analyzed by the respective receiver operating characteristic (ROC) curve where 1-specificity and sensitivity are plotted against each other as a function of the cutoff of the underlying classification probability. After computing each ROC curve, we calculated the respective area under the ROC curve (AUC) and its 95% confidence interval to determine the accuracy of the classification irrespective of the cut-off used. In general, an AUC=0.50 is indicative of a complete random classification of the individuals while AUC=1.00 implies that the constructed classifier perfectly predicts the true class membership of each individual.

We performed further antibody-wide association analyses related to the following comparisons (or classification exercises): (i) healthy controls versus all ME/CFS patients; (ii) healthy controls versus ME/CFS patients with an infectious trigger; (iii) healthy controls versus ME/CFS patients with a noninfectious or unknown trigger; and (iv) ME/CFS patients with an infectious trigger versus the remaining ME/CFS patients. In each association analysis, we first estimated three regression models: logistic model, probit model, and complementary log-log model. In these models, the disease status was the outcome variable, and age and gender were the respective covariates. To determine the best link function for the outcome variable, we selected the model with the lowest Akaike’s information criterion (AIC). For the best link function (“the null model”), we estimated the respective ROC and its AUC as described above. We fitted five different logistic models, including the main effects and all possible interaction terms among age, gender, and the antibody response under analysis: (i) a model with main effects only and no interaction terms; (ii) a model with an interaction term between age and the antibody response; (iii) a model with an interaction term between gender and the antibody response; (iv) a model with two interaction terms between age and the antibody response and between gender and the antibody response; (v) a model with all possible two-way and three-way interaction among age, gender, and the antibody response. We compared each of these models with the null one using Wilks’ likelihood ratio test, where low p-values provide evidence for these models, including effects of an antibody response. We reported the minimum p-value obtained from these model comparisons. Finally, we adjusted the minimum p-values of each analysis. This adjustment was made using the Benjamini-Yekutieli procedure ensuring a global false discovery rate (FDR) of 5% under the assumption of dependent tests (31). In this analysis, adjusted p-values <0.05 indicated statistically significant results.

To filter out redundant antibody responses, we pooled all the significant antibody responses in a single logistic model. The effect and interaction terms of these antibody responses were defined according to the most significant model obtained in the previous stage of analysis. We performed a backward stepwise model selection. The resulting model was finally evaluated in terms of predictive performance using ROC analysis as described above.

The above analysis was primarily done for the whole data set irrespective of the ME/CFS subgroups. We repeated the same analysis to compare each subgroup of ME/CFS patients (with infectious and noninfectious or unknown disease trigger) with the healthy controls. Finally, we repeated the analysis to compare the two subgroups of ME/CFS patients.

### Statistical software

The statistical analysis was performed in the R software version 4.0.3 with core functions and the following packages: MASS v7.3-56 to perform stepwise model selection (32), pROC v1.18.0 to estimate the ROC curve and the respective AUC (33), OptimalCutpoints v1.1-5 to estimate the optimal cutoff and the associated sensitivity/specificity (34). The full reproducible code is are freely available from NS or JM upon request.

### Ethical approval

All individuals gave written informed consent to participate in the study. The study was approved by the Ethics Committee of Charité Universitatsmedizin Berlin in accordance with the 1964 Declaration of Helsinki and its later amendments (23).

## RESULTS

### Principal component and linear discriminant analyses

We first performed a PCA to discriminate patients with ME/CFS and their subgroups from healthy controls (Figures 1A, 1B, and 1C). A similar analysis was done for discriminating patients with an infectious trigger from patients with noninfectious or unknown trigger (Figure 1D).

**Figure 1.**
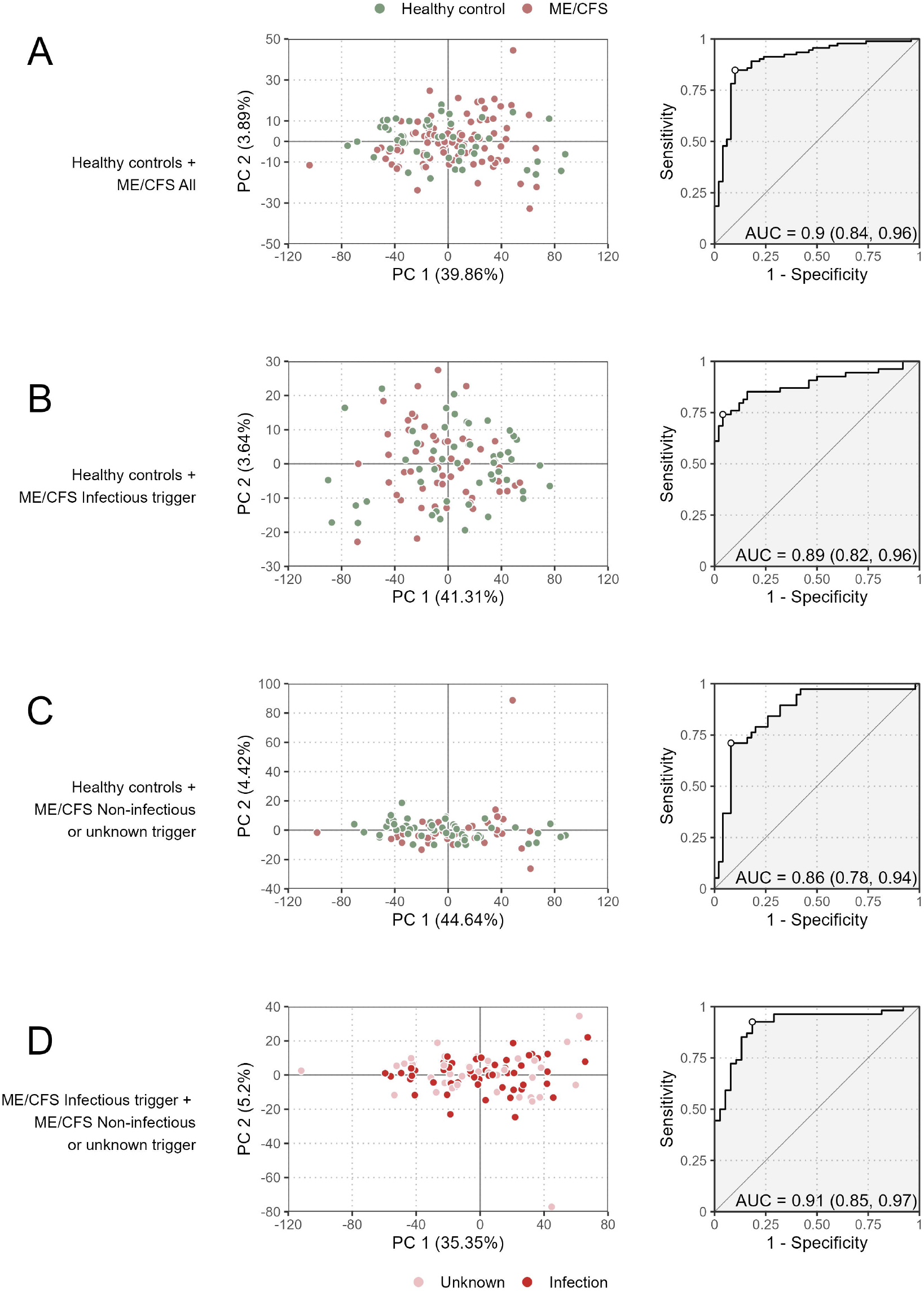
Preliminary multivariate analysis of the data. Scatterplots of the first two principal components (left plots) and the ROC curve and its AUC of the respective LDA (right plots) when comparing all ME/CFS patients to healthy controls (A), ME/CFS patients with an infectious trigger to healthy controls (B), ME/CFS patients with a noninfectious or unknown trigger to healthy controls (C), and ME/CFS patients with an infectious trigger to the remaining patients (D). The percentage of the variance explained by each principal component is shown within brackets in each axis.

The proportion of variance explained by the first principal component varied from 35.4% (Figure 1D) to 44.6% (Figure 1C) referring to the comparisons between the two subgroups of ME/CFS patients, and between healthy controls and patients with non-infectious or unknown disease trigger, respectively. These high estimates associated with the first principal component suggested that the antibody levels were correlated with each other. This interpretation was confirmed by determining the distributions of Spearman’s correlation coefficient between all possible pairs of antibodies using data from each study group (Supplementary Figure 1). In particular, the antibody levels were positively correlated with each other with median correlation estimates of 0.56, 0.56, 0.40, and 0.48 for healthy controls, all ME/CFS patients, ME/CFS patients with an infectious disease trigger, and the remaining ME/CFS patients, respectively. Interestingly, the median correlation estimate was decreased in ME/CFS patients with an infectious trigger when compared to other study groups. This finding suggested that the production of the antibodies against the EBV-derived antigens could be reduced in these patients when compared to healthy controls or patients with non-infectious or unknown disease trigger.

The visualization of the first two components did not reveal a clear discrimination between healthy controls and ME/CFS patients (or their subgroups). To improve this analysis, we then performed different LDAs in search of a linear combination of the antibody measurements that could be used for disease diagnosis. The performance of the constructed classifiers ranged from 0.86 (Figure 1C) to 0.91 (Figure 1D) referring to the classification of healthy controls and ME/CFS patients with noninfectious or unknown disease trigger and the classification of the two subgroups of ME/CFS patients, respectively. Therefore, the results of this analysis indicate that the obtained data could discriminate different study groups.

### Antibody-wide association analysis

The next step of the analysis was to identify specific antibody responses that could be used to discriminate the different study groups. With this purpose, we first determined the best “null” model among the logistic, probit, and complementary log-log models. All of them included age and gender and their interaction as covariates for each comparison between any two study groups (Supplementary Table 2). The best ‘‘null’ models were the following: (i) (i) complementary log-log -comparison between healthy controls and all the ME/CFS patients (AUC= 0.574; 95% CI=(0.475;0.672)); (ii) probit – comparison between healthy controls and ME/CFS patients with an infectious trigger (AUC=0.606; 95% CI=(0.496;0.715)); (iii) complementary log-log - comparison between healthy controls and ME/CFS patients with a noninfectious or unknown trigger (AUC= 0.556; 95% CI=(0.429;0.683)); and (iv) logit – comparison between the two subgroups of ME/CFS groups (AUC=0.596; 95% CI=(0.471;0.720)). The 95% confidence interval for the AUC of these null models included 0.50 and therefore, the respective predicted classification was consistent with a random guess. Such a result was in agreement with the age and gender matching between different study groups and healthy controls (Table 1).

We performed further antibody-wide association analyses controlling for a global FDR of 5%. The comparison between healthy controls and all ME/CFS patients did not identify any significant antibody associations with the disease (Figure 2A). The top 5 antibodies, although not statistically significant, were EBNA6_0066, BLRF2_0005, EBNA4_0392, EBNA4_0497, and EBNA4_0529 (adjusted p-values = 0.181, 0.326, 0.326, 0.326, and 0.326, respectively).

**Figure 2.**
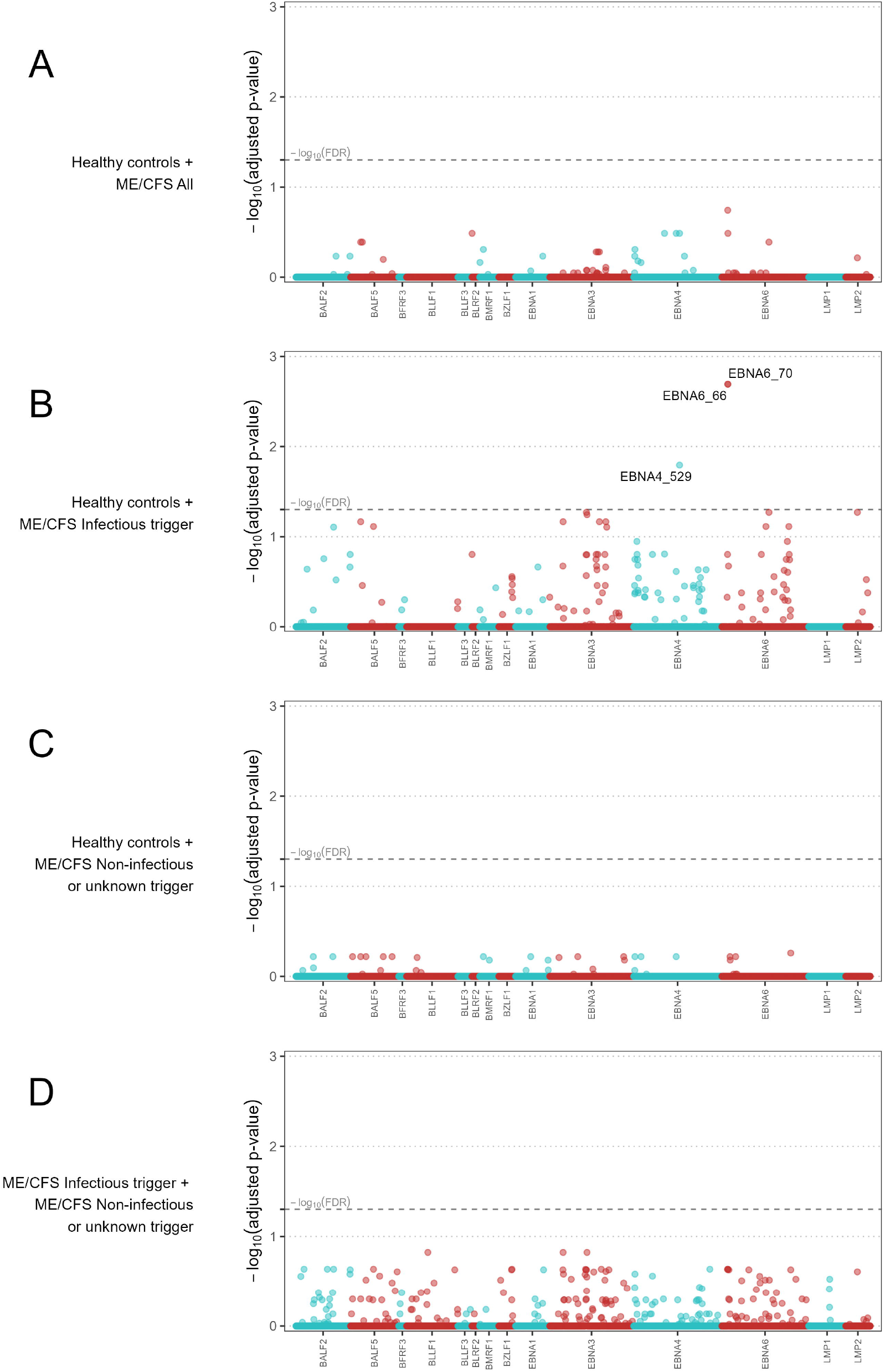
Antibody-wide association analyses concerning comparison where x-axis comprises the association of each antibody and y-axis represents the -log_10_(adjusted p-value). In the x-axis, the antibodies were ordered alphabetically first by the protein name and then by the starting point of the antigen within a given protein. Adjusted p-values were adjusted according to the Benjamini-Yekutieli procedure for a global FDR of 5% under the assumption of dependent data. Dashed line represents the threshold for statistical significance (i.e., -log_10_(FDR=0.05)) and -log_10_(adjusted p-values) > 1.30 were considered statistically significant.

When the comparison was limited to healthy controls and ME/CFS patients with an infectious trigger, we identified three significant antibodies related to the following antigens (Figure 2B): EBNA6_0066, EBNA6_0070, and EBNA4_0529 (adjusted p-values=0.005, 0.005, 0.038, respectively). The first two antigens were shared between AG876, B95-8, and GD1 strains, while the third one was derived from the B95-8 strain. We compared ME/CFS patients with noninfectious or unknown disease trigger to healthy controls, and found no significant differences in antibody responses (Figure 2C). The same finding was obtained when we compared the two subgroups of ME/CFS patients (Figure 2D). The top 5 antibodies related to these analyses can be found in Supplementary Table 3.

### Analysis of candidate antigens for classifying ME/CFS patients with an infectious trigger

We then analyzed in detail the impact of the antibody levels against the three candidate antigens on the classification of ME/CFS patients with an infectious trigger. Antibody levels were increased in this subgroup of ME/CFS patients when compared to healthy controls (Figure 3A). The same evidence could not be found when comparing all ME/CFS patients to healthy controls (Figure 3A). Data related to EBNA4_0529, EBNA6_0066 and EBNA6_0070 were significantly correlated with each other (Spearman’s correlation coefficients higher than 0.58; Figure 3B). The correlation between the levels of antibodies against EBNA6_0066 and EBNA6_0070 could be explained by the fact that these two peptides are 15-mers overlapping 11 amino acids with each other (23). In contrast, it was unclear why the levels of antibodies against EBNA4_0529 and EBNA6_0066 were highly correlated (Spearman’s correlation coefficient = 0.79), considering that these antigens did not share a high sequence homology (Figure 3C).

**Figure 3.**
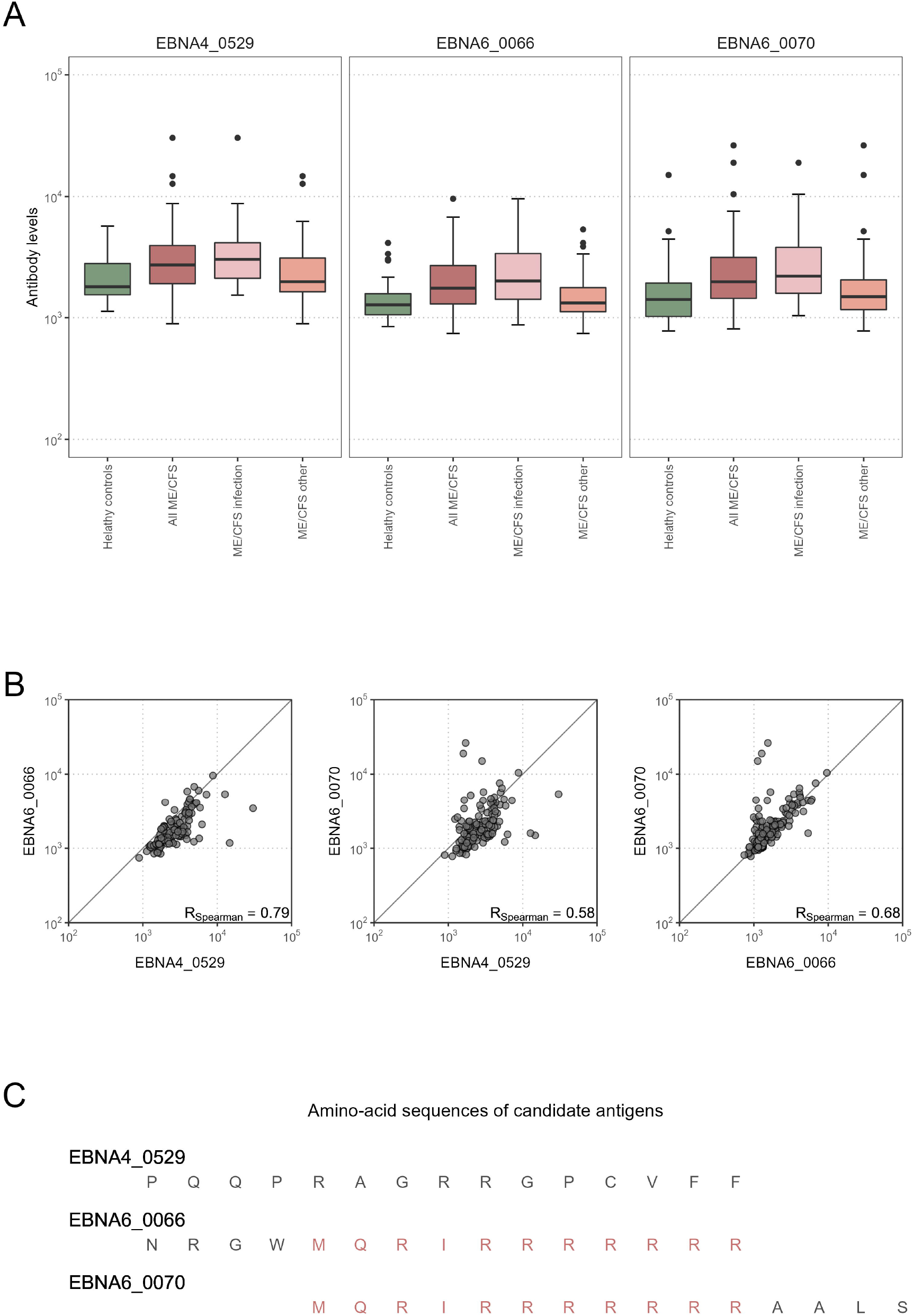
Statistical analysis of the antibody levels related to EBNA4_0529, EBNA6_0066, and EBNA6_0070. **A**. Boxplots of the data per study group. **B**. Scatterplots and the respectively Spearman’s correlation coefficients (R) in the whole dataset. **C**. Amino acid sequences of EBNA4_0529, EBNA6_0066, and EBNA6_0070.

Given the high correlation between antibody levels related to these antigens, some statistical redundancy was expected when using their data for patients’ classification purpose. This redundancy was confirmed when the three candidate antibodies were included as covariates in the same model. A stepwise variable selection procedure led to the exclusion of the antibody levels related to EBNA6_0066 from the final classification model.

The final model included the main effects of antibodies to EBNA4_0529 and EBNA6_0070 and the two-way interaction of the latter with age and gender (Table 2). On the one hand, the log_10_-levels of antibodies related to EBNA4 increased the probability of being a patient (coefficient estimate = 2.25, Standard error=1.09). In particular, the odds of being a patient were estimated to increase ∼9.5 (e^2.25^) times per fold-change in the levels of these antibodies. On the other hand, the effects of antibody levels related to EBNA6_0070 on the probability of an individual being an ME/CFS patients were not so trivial to ascertain (Figure 4A). In particular, women with high EBNA6_0070 antibody levels showed an increasing estimated probability of being a patient with increasing age. In contrast, the probability profile of being patient was different in men. In that case, younger men with low EBNA6_0070 antibody levels or older men with high EBNA6 antibody levels had a higher probability of being a patient.

**Table 2.**
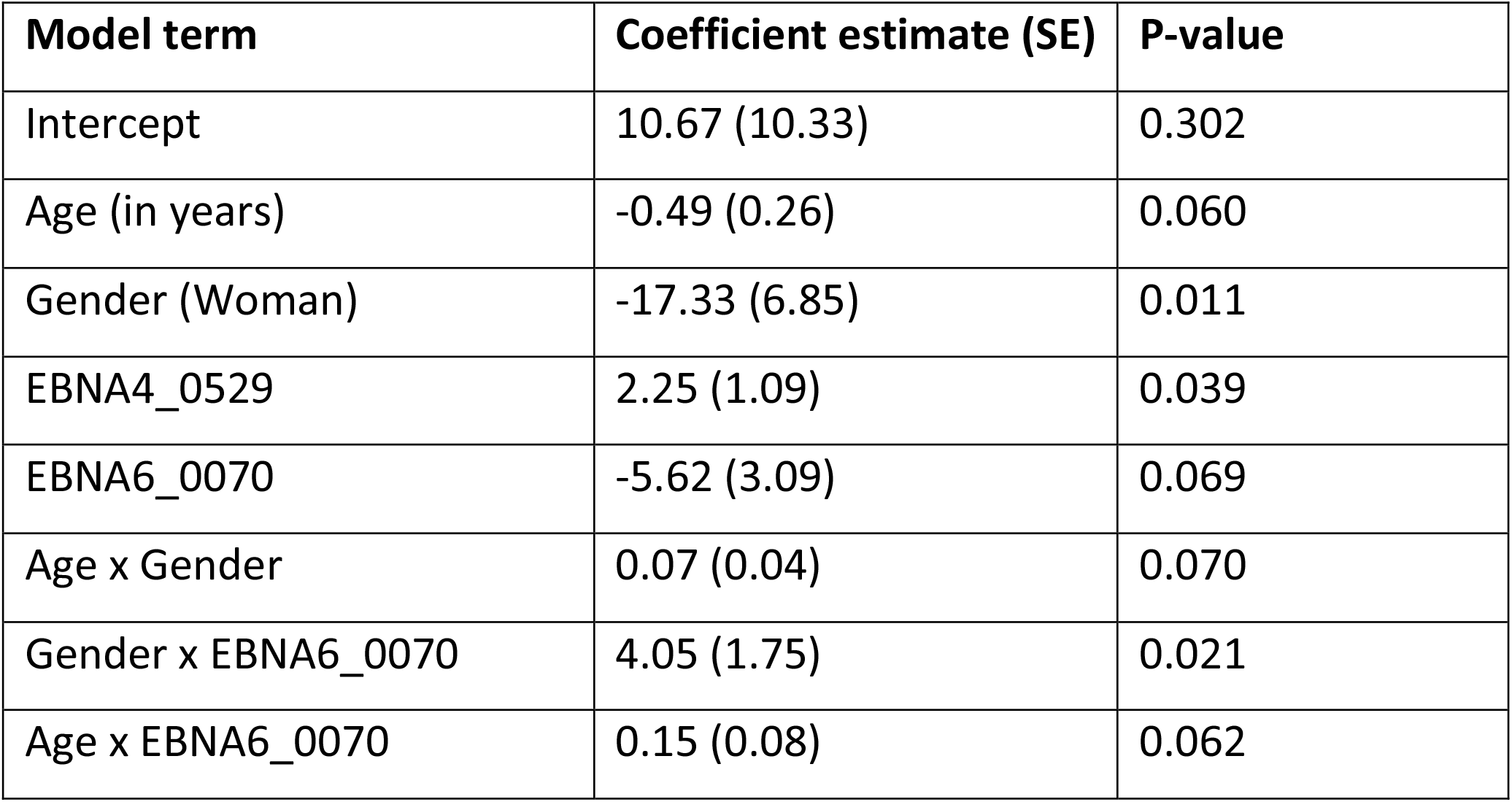
Estimates of the final complementary log-log model to discriminate patients with an infectious disease trigger from healthy controls.

**Figure 4.**
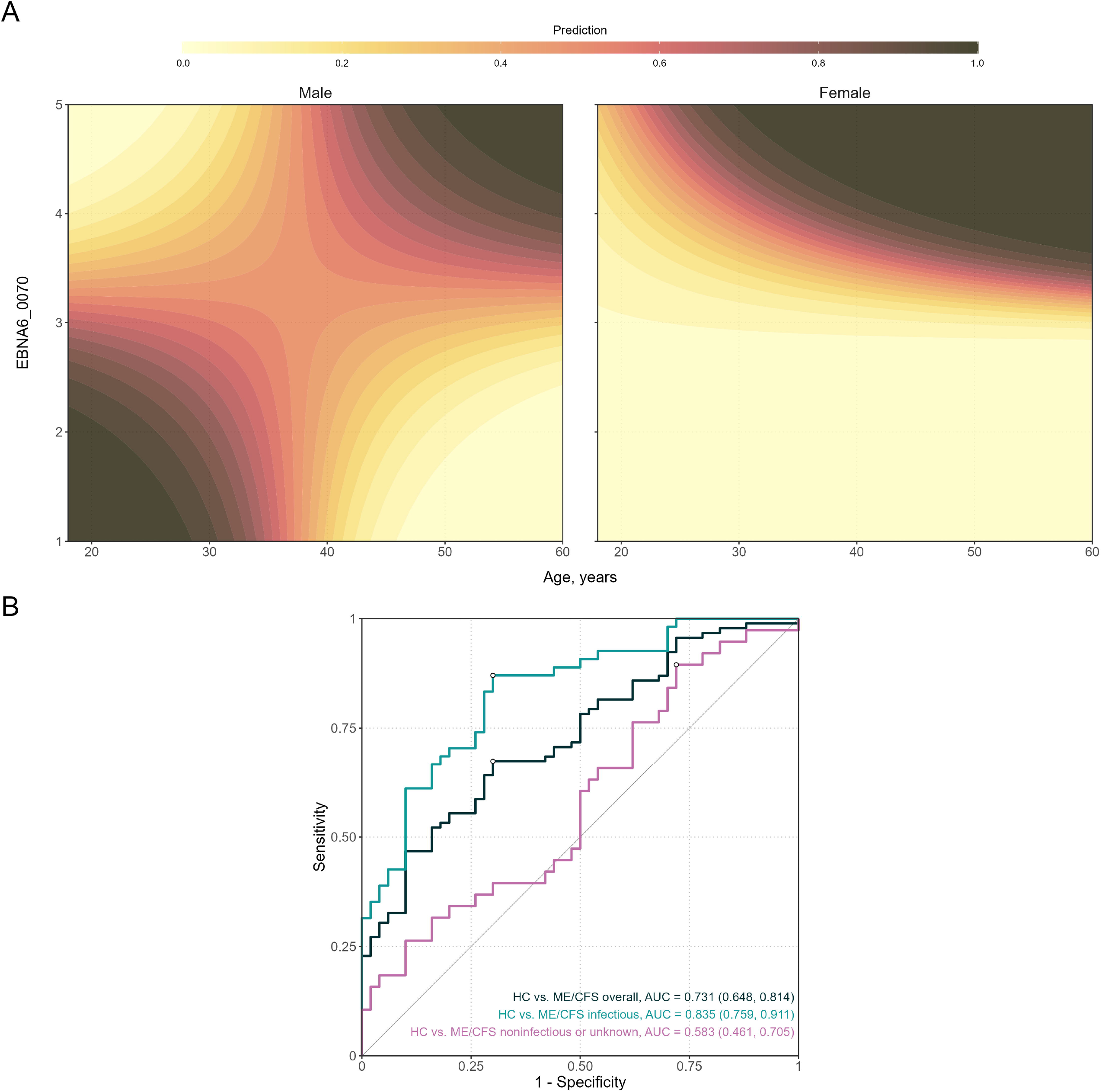
Analysis of the final classification model for predicting ME/CFS patients with infectious trigger when compared to healthy controls. **A**. Contour plots of the probability of being a ME/CFS patient as a function of age and EBNA6_0070 antibody levels, for men and women, respectively. The prediction values were calculated by fixing log_10_(EBNA4_0529) at the respective mean value. **B**. ROC curves and the respective AUC (95% confidence interval shown within brackets) when using the model to compare different groups of ME/CFS patients to healthy controls.

The AUC of the classification predicted by the final model was estimated at 0.835 with a 95% CI=(0.759;0.911) (Figure 4B). This estimate suggested that the combination of these two antibodies together with age and gender could be used for the diagnosis of patients with an infectious trigger. The optimal sensitivity and specificity were estimated at 0.833 and 0.720, respectively. Therefore, ME/CFS patients were better discriminated by this model than healthy controls.

When the same classification model was applied to the whole cohort of ME/CFS patients, the AUC decreased to 0.731 with a 95% CI=(0.648, 0.814). This could be explained by the cohort of patients with a non-infectious or unknown trigger in which the performance of the classification model was close to a random guess (AUC=0.583; 95% CI=(0.461;0.705)).

## DISCUSSION

This study, based on previously published data, aimed to discover EBV-derived antigens that could elicit distinct antibody responses in ME/CFS patients when compared to healthy controls using previously published data. The key finding was the identification of two candidate antigens inducing increased antibody responses in ME/CFS patients with an infectious trigger. The high sensitivity and specificity of our classification model including these antibodies suggest their potential for diagnosis of this subgroup of affected individuals. For ME/CFS patients without an infectious trigger, we could not find any antigens causing antibody responses that could be used for diagnostic purposes. This finding is in agreement with an extensive serological investigation of different herpesviruses in ME/CFS patients (29). This negative finding supports the hypothesis that EBV plays a role in the group of ME/CFS patients with an infectious trigger. In a subset of patients, infectious mononucleosis caused by primary EBV infection can be documented as a trigger (10). In many others, no infection with a specific pathogen could be associated with disease onset (5). A tempting hypothesis from our finding is that EBV reactivation which can occur during other infections may play until now an underestimated role in triggering ME/CFS. In line with this concept, a recent study showed that EBV reactivation during COVID-19 is a risk factor for Post COVID Syndrome which also includes ME/CFS (35). Alternatively, the responses to the EBNA6 peptides are due to a cross-reactivity to other pathogens, as outlined below.

Other findings of this study pointed to three key challenges associated with the discovery of a biomarker. Firstly, it is difficult to identify a disease-specific biomarker for all the ME/CFS patients. Thus, given the heterogeneous nature of ME/CFS, it is pivotal to stratify patients adequately (30), based on age, gender, and disease trigger for biomarker discovery (27). In this regard, the identification of antibody patterns specific to ME/CFS patients with an infectious trigger was in agreement with other studies where significant results could be found for subgroup of ME/CFS patients with infectious triggers (10,36,37). However, given the vast number of infectious agents associated with ME/CFS (5,38), it is worth noting that this subgroup of patients could be further subdivided according to the nature of the causative infection. In this regard, the data about the infectious agents that could have initiated ME/CFS are either inconclusive or simply based on self-reported history in most patients, as demonstrated by the data from the United Kingdom ME/CFS Biobank, where only a minority of patients had their infection confirmed with the lab test (10). Secondly, the final classification model included non-trivial statistical interactions of antibodies against EBNA6_0070 with both age and gender. This finding implies that significant interactions between candidate biomarkers and confounding factors might be overlooked by analysts or, even when tested, they are likely to be discarded due to the small sample sizes to detect them. The presence of these interactions might be yet another factor that contributes to the lack of reproducibility between biomarker studies on ME/CFS. A proposed strategy to overcome this limitation is to conduct more advanced statistical analyses including the application of machine learning techniques which intrinsically consider the complexity of a large set of clinical and biological data, as demonstrated in drug discovery (39). Thirdly, the interaction between the candidate antibodies against EBNA6_0070 and gender implied a remarkable distinct antibody signature between male and female patients. Again, this finding is in line with gender differences in immunity to viral infection (40). In particular, men have typically lower antibody responses when vaccinated and are more susceptible to infections than women (41). In this regard, our study suggested that the higher probability of younger man being an ME/CFS patient is associated with lower levels of antibodies against the antigen EBNA6_0070. In contrast, female and male patients seemed to be at higher risk with higher antibodies at increasing age suggesting that at least a subset develop these antibody responses later in life. An implication of having a different antibody profiling between men and women is that analysis of each gender should be performed separately. At the same time, it is important to note that epidemiological data on ME/CFS suggested approximately a disease ratio of three women to one man (42–44). Therefore, if gender is an important stratification factor for biomarker discovery, studies should be designed towards a more balanced gender ratio. Similar sample sizes between male and female cohorts ensure comparable statistical power when analysing data from each sex separately.

Both EBNA4_0529 and EBNA6_0070 antigens are derived from proteins whose genetic expression typically occurs during the EBV type III latency. Therefore, the acquisition of the respective antibodies might have occurred during initial B-cell transformation and immortalization. It could also be acquired slowly over time, given that the type III latency pattern can be detected sporadically in lymphoid follicles where EBV-infected B cells can proliferate and mimic a germinal center reaction program (45). We can hypothesize from our data that both male and female patients developing higher antibody responses against this antigen later in live are at increased risk of developing ME/CFS suggesting that reactivation of EBV plays a role. In male patients a subgroup with lower EBNA6 antibodies early in live is at risk of developing ME/CFS, too. Using the recent analytical framework of ME/CFS natural progression (46), antibodies against these antigens are more likely to be biomarkers of patients suffering from ME/CFS more than two years of disease rather than the ones either in prodromal period or at early stages in line with our findings. Based on that assumption, these antibodies seemed more appropriate for diagnosing putative patients with delayed disease diagnosis rather than early suspected cases. However, it is known that the delay of ME/CFS diagnosis is a recurrent problem in the clinic (8,47). As such, we anticipate a higher utility of these antibodies when redeployed to real-world screening. Another practical implication of using these antibodies as biomarkers is the possibility of developing routine ELISA kits that can be standardized across different laboratories and easily scalable for large population screenings. Notwithstanding these promising practical expectations, it is important to emphasize that past studies also suggested potential disease biomarkers (27) and, therefore, it is imperative to replicate the findings of this study with different cohorts of patients.

An interesting observation is that both EBNA6_0066 and EBNA6_0070 contain an arginine-reach sequence. Such a sequence has homologies with putative epitopes from several human proteins (48). Such homologies suggest a potential molecular mimicry between the viral and human antigens. Molecular mimicry can trigger deleterious autoimmune responses as hypothesized for ME/CFS pathogenesis (38,49). Molecular mimicry between human and microbial antigens has been also hypothesized for several autoimmune diseases (50), such as multiple sclerosis and rheumatoid arthritis, and Post COVID syndrome, whose patients share similar symptoms with ME/CFS ones (19,51–53). Interestingly, T cell clones recognizing such arginine-repeat sequences were isolated from a patient with multiple sclerosis supporting our concept of epitope mimicry (48). Finally, arginine-repeat sequences are found in various other pathogens including enteroviruses and human papillomavirus which are also triggers of ME/CFS (5).

Further we can hypothesize that peptides highly enriched in arginine residues might be particularly susceptible to citrullination, in which arginine residues are post-translationally converted to citrulline. These post-translational modifications occur during cell death under normal physiological conditions. However, under chronic inflammation, the accumulation of citrullinated (auto)antigens in inflamed sites might lead to deleterious autoimmune responses, thus, promoting the onset of different autoimmune diseases (54). A potential cross-reactivity between microbial and citrullinated human antigens could also be a mechanism by which an autoimmune disease can be triggered. In rheumatoid arthritis antibodies against EBNA-1 peptides were shown to cross-react with denatured collagen and keratin (55). However, in the present study, we could not find any antibodies against EBNA-1-derived peptides to be associated with ME/CFS. Interestingly, the serum levels of citrulline were reported to be elevated in ME/CFS patients when compared to healthy controls (56). However, another study could not confirm this finding, but instead provided evidence for increased plasma levels of arginine residues (57). Another source of antigen modification is the process of generating new and more immunogenic epitopes from ubiquitous molecules upon oxidative and nitrosative stress. In ME/CFS, IgM antibodies against several of these neoepitopes, including NO-Arginine, were increased in patients (58). In all these possible scenarios, it is imperative to investigate the stability of this candidate biomarker antigen to post-translational modifications that could be occurred and eventually increased during the disease course.

In conclusion, this study identified two candidate antigens whose antibodies could be used to identify ME/CFS patients with an infectious trigger. To strengthen our findings, two other cohorts of patients are currently studied, including the well-characterized ME/CFS patients with different disease triggers and healthy controls from the United Kingdom ME/CFS biobank (10).

## Data Availability

Antibody data are available online at: https://doi.org/10.1371/journal.pone.0179124.s001. The remaining data are available upon request to Prof. Carmen Scheibenbogen.

https://doi.org/10.1371/journal.pone.0179124.s001

## Authors’ contributions

NS and CS conceived this research. NS, JM, ADG, and AF performed the data analysis. FS, UB, and CS collected and provided the data. All authors interpreted and discussed the results. NS wrote the paper. All authors have read, revised, and approved the final version of the manuscript.

## REFERENCES

1. Houen G, Trier NH. Epstein-Barr Virus and Systemic Autoimmune Diseases. Front Immunol (2020) 11:587380. doi:10.3389/fimmu.2020.587380

2. Shannon-Lowe C, Rickinson AB, Bell AI. Epstein–Barr virus-associated lymphomas. Philos Trans R Soc B Biol Sci (2017) 372:20160271. doi:10.1098/rstb.2016.0271

3. Bjornevik K, Cortese M, Healy BC, Kuhle J, Mina MJ, Leng Y, Elledge SJ, Niebuhr DW, Scher AI, Munger KL, et al. Longitudinal analysis reveals high prevalence of Epstein-Barr virus associated with multiple sclerosis. Science (80-) (2022) 375:296–301. doi:10.1126/science.abj8222

4. Koo D. Chronic fatigue syndrome. A critical appraisal of the role of Epstein-Barr virus. West J Med (1989) 150:590–596.

5. Rasa S, Nora-Krukle Z, Henning N, Eliassen E, Shikova E, Harrer T, Scheibenbogen C, Murovska M, Prusty BK. Chronic viral infections in myalgic encephalomyelitis/chronic fatigue syndrome (ME/CFS). J Transl Med (2018) 16:268. doi:10.1186/s12967-018-1644-y

6. Ruiz-Pablos M, Paiva B, Montero-Mateo R, Garcia N, Zabaleta A. Epstein-Barr Virus and the Origin of Myalgic Encephalomyelitis or Chronic Fatigue Syndrome. Front Immunol (2021) 12:656797. doi:10.3389/fimmu.2021.656797

7. Rivera MC, Mastronardi C, Silva-Aldana CT, Arcos-Burgos M, Lidbury BA. Myalgic encephalomyelitis/chronic fatigue syndrome: A comprehensive review. Diagnostics (2019) 9:91. doi:10.3390/diagnostics9030091

8. Bateman L, Bested AC, Bonilla HF, Chheda B V., Chu L, Curtin JM, Dempsey TT, Dimmock ME, Dowell TG, Felsenstein D, et al. Myalgic Encephalomyelitis/Chronic Fatigue Syndrome: Essentials of Diagnosis and Management. Mayo Clin Proc (2021) 96:2861–2878. doi:10.1016/j.mayocp.2021.07.004

9. Hickie I, Davenport T, Wakefield D, Vollmer-Conna U, Cameron B, Vernon SD, Reeves WC, Lloyd A. Post-infective and chronic fatigue syndromes precipitated by viral and non-viral pathogens: prospective cohort study. BMJ (2006) 333:575. doi:10.1136/bmj.38933.585764.AE

10. Domingues TD, Grabowska AD, Lee JS, Ameijeiras-Alonso J, Westermeier F, Scheibenbogen C, Cliff JM, Nacul L, Lacerda EM, Mouriño H, et al. Herpesviruses Serology Distinguishes Different Subgroups of Patients From the United Kingdom Myalgic Encephalomyelitis/Chronic Fatigue Syndrome Biobank. Front Med (2021) 8:686736. doi:10.3389/fmed.2021.686736

11. Shikova E, Reshkova V, Kumanova A, Raleva S, Alexandrova D, Capo N, Murovska M, on B. Cytomegalovirus, Epstein-Barr virus, and human herpesvirus-6 infections in patients with myalgic encephalomyelitis/chronic fatigue syndrome. J Med Virol (2020) 92:3682. doi:10.1002/jmv.25744

12. Lee JS, Lacerda EM, Nacul L, Kingdon CC, Norris J, O’Boyle S, Roberts C h., Palla L, Riley EM, Cliff JM. Salivary DNA Loads for Human Herpesviruses 6 and 7 Are Correlated With Disease Phenotype in Myalgic Encephalomyelitis/Chronic Fatigue Syndrome. Front Med (2021) 8:1129. doi:10.3389/fmed.2021.656692

13. Kerr JR. Epstein-Barr Virus Induced Gene-2 Upregulation Identifies a Particular Subtype of Chronic Fatigue Syndrome/Myalgic Encephalomyelitis. Front Pediatr (2019) 7:59. doi:10.3389/fped.2019.00059

14. Lerner AM, Ariza ME, Williams M, Jason L, Beqaj S, Fitzgerald JT, Lemeshow S, Glaser R. Antibody to Epstein-Barr virus deoxyuridine triphosphate nucleotidohydrolase and deoxyribonucleotide polymerase in a chronic fatigue syndrome subset. PLoS One (2012) 7:e47891. doi:10.1371/journal.pone.0047891

15. Loebel M, Strohschein K, Giannini C, Koelsch U, Bauer S, Doebis C, Thomas S, Unterwalder N, Von Baehr V, Reinke P, et al. Deficient EBV-specific B-and T-cell response in patients with Chronic Fatigue Syndrome. PLoS One (2014) 9:e85387. doi:10.1371/journal.pone.0085387

16. Wang J, Jelcic I, Mühlenbruch L, Haunerdinger V, Toussaint NC, Zhao Y, Cruciani C, Faigle W, Naghavian R, Foege M, et al. HLA-DR15 Molecules Jointly Shape an Autoreactive T Cell Repertoire in Multiple Sclerosis. Cell (2020) 183:1264–1281.e20. doi:10.1016/j.cell.2020.09.054

17. Malato J, Graça L, Nacul L, Lacerda E, Sepúlveda N. Statistical challenges of investigating a disease with a complex diagnosis. medRxiv (2021)2021.03.19.21253905. doi:10.1101/2021.03.19.21253905

18. Morris G, Maes M. Myalgic encephalomyelitis/chronic fatigue syndrome and encephalomyelitis disseminata/multiple sclerosis show remarkable levels of similarity in phenomenology and neuroimmune characteristics. BMC Med (2013) 11:205. doi:10.1186/1741-7015-11-205

19. Gaber TAZK, Oo WW, Ringrose H. Multiple Sclerosis/Chronic Fatigue Syndrome overlap: When two common disorders collide. NeuroRehabilitation (2014) 35:529–534. doi:10.3233/NRE-141146

20. Wucherpfennig KW, Strominger JL. Molecular mimicry in T cell-mediated autoimmunity: Viral peptides activate human T cell clones specific for myelin basic protein. Cell (1995) 80:695–705. doi:10.1016/0092-8674(95)90348-8

21. Holmøy T, Kvale EØ, Vartdal F. Cerebrospinal fluid CD4+ T cells from a multiple sclerosis patient cross-recognize Epstein-Barr virus and myelin basic protein. J NeuroVirology 2004 105 (2004) 10:278–283. doi:10.1080/13550280490499524

22. Lünemann JD, Jelčić I, Roberts S, Lutterotti A, Tackenberg B, Martin R, Münz C. EBNA1-specific T cells from patients with multiple sclerosis cross react with myelin antigens and co-produce IFN-γ and IL-2. J Exp Med (2008) 205:1763–1773. doi:10.1084/jem.20072397

23. Loebel M, Eckey M, Sotzny F, Hahn E, Bauer S, Grabowski P, Zerweck J, Holenya P, Hanitsch LG, Wittke K, et al. Serological profiling of the EBV immune response in Chronic Fatigue Syndrome using a peptide microarray. PLoS One (2017) 12:e0179124. doi:10.1371/journal.pone.0179124

24. Tengvall K, Huang J, Hellström C, Kammer P, Biström M, Ayoglu B, Bomfim IL, Stridh P, Butt J, Brenner N, et al. Molecular mimicry between Anoctamin 2 and Epstein-Barr virus nuclear antigen 1 associates with multiple sclerosis risk. Proc Natl Acad Sci U S A (2019) 116:16955–16960. doi:10.1073/pnas.1902623116

25. Sepúlveda N. Impact of genetic variation on the molecular mimicry between Anoctamin-2 and Epstein-Barr virus nuclear antigen 1 in Multiple Sclerosis. Immunol Lett (2021) 238:29–31. doi:10.1016/j.imlet.2021.07.007

26. Sepúlveda N, Carneiro J, Lacerda E, Nacul L. Myalgic Encephalomyelitis/Chronic Fatigue Syndrome as a Hyper-Regulated Immune System Driven by an Interplay Between Regulatory T Cells and Chronic Human Herpesvirus Infections. Front Immunol (2019) 10:2684. doi:10.3389/fimmu.2019.02684

27. Scheibenbogen C, Freitag H, Blanco J, Capelli E, Lacerda E, Authier J, Meeus M, Castro Marrero J, Nora-Krukle Z, Oltra E, et al. The European ME/CFS Biomarker Landscape project: An initiative of the European network EUROMENE. J Transl Med (2017) 15:162. doi:10.1186/s12967-017-1263-z

28. Cliff JM, King EC, Lee JS, Sepúlveda N, Wolf AS, Kingdon C, Bowman E, Dockrell HM, Nacul L, Lacerda E, et al. Cellular immune function in myalgic encephalomyelitis/chronic fatigue syndrome (ME/CFS). Front Immunol (2019) 10:796. doi:10.3389/fimmu.2019.00796

29. Blomberg J, Rizwan M, Böhlin-Wiener A, Elfaitouri A, Julin P, Zachrisson O, Rosén A, Gottfries CG. Antibodies to human herpesviruses in myalgic encephalomyelitis/chronic fatigue syndrome patients. Front Immunol (2019) 10:1946. doi:10.3389/fimmu.2019.01946

30. Jason LA, Corradi K, Torres-Harding S, Taylor RR, King C. Chronic fatigue syndrome: The need for subtypes. Neuropsychol Rev (2005) 15:29–58. doi:10.1007/s11065-005-3588-2

31. Benjamini Y, Yekutieli D. The control of the false discovery rate in multiple testing under dependency. Ann Stat (2001) 29:1165–1188. doi:10.1214/aos/1013699998

32. Venables WN, Ripley BD. Modern Applied Statistics with S. Fourth. New York: Springer (2002). Available at: https://www.stats.ox.ac.uk/pub/MASS4/ [Accessed May 28, 2021]

33. Robin X, Turck N, Hainard A, Tiberti N, Lisacek F, Sanchez JC, Müller M. pROC: An open-source package for R and S+ to analyze and compare ROC curves. BMC Bioinformatics (2011) 12:1–8. doi:10.1186/1471-2105-12-77

34. López-Ratón M, Rodríguez-Álvarez MX, Cadarso-Suárez C, Gude-Sampedro F. Optimalcutpoints: An R package for selecting optimal cutpoints in diagnostic tests. J Stat Softw (2014) 61:1–36. doi:10.18637/jss.v061.i08

35. Su Y, Yuan D, Chen DG, Ng RH, Wang K, Choi J, Li S, Hong S, Zhang R, Xie J, et al. Multiple early factors anticipate post-acute COVID-19 sequelae. Cell (2022) 185:881–895.e20. doi:10.1016/j.cell.2022.01.014

36. Steiner S, Becker SC, Hartwig J, Sotzny F, Lorenz S, Bauer S, Löbel M, Stittrich AB, Grabowski P, Scheibenbogen C. Autoimmunity-Related Risk Variants in PTPN22 and CTLA4 Are Associated With ME/CFS With Infectious Onset. Front Immunol (2020) 11:578. doi:10.3389/fimmu.2020.00578

37. Szklarski M, Freitag H, Lorenz S, Becker SC, Sotzny F, Bauer S, Hartwig J, Heidecke H, Wittke K, Kedor C, et al. Delineating the Association Between Soluble CD26 and Autoantibodies Against G-Protein Coupled Receptors, Immunological and Cardiovascular Parameters Identifies Distinct Patterns in Post-Infectious vs. Non-Infection-Triggered Myalgic Encephalomyelitis/Chro. Front Immunol (2021) 12:1077. doi:10.3389/fimmu.2021.644548

38. Blomberg J, Gottfries CG, Elfaitouri A, Rizwan M, Rosén A. Infection elicited autoimmunity and Myalgic encephalomyelitis/chronic fatigue syndrome: An explanatory model. Front Immunol (2018) 9:229. doi:10.3389/fimmu.2018.00229

39. Gupta R, Srivastava D, Sahu M, Tiwari S, Ambasta RK, Kumar P. Artificial intelligence to deep learning: machine intelligence approach for drug discovery. Mol Divers (2021) 25:1315–1360. doi:10.1007/S11030-021-10217-3

40. Jacobsen H, Klein SL. Sex Differences in Immunity to Viral Infections. Front Immunol (2021) 12:3483. doi:10.3389/fimmu.2021.720952

41. Aaby P, Benn CS, Flanagan KL, Klein SL, Kollmann TR, Lynn DJ, Shann F. The non-specific and sex-differential effects of vaccines. Nat Rev Immunol (2020) 20:464–470. doi:10.1038/S41577-020-0338-X

42. Chu L, Valencia IJ, Garvert DW, Montoya JG. Onset patterns and course of myalgic encephalomyelitis/chronic fatigue syndrome. Front Pediatr (2019) 7:12. doi:10.3389/fped.2019.00012

43. Johnston SC, Staines DR, Marshall-Gradisnik SM. Epidemiological characteristics of chronic fatigue syndrome/myalgic encephalomyelitis in Australian patients. Clin Epidemiol (2016) 8:97–107. doi:10.2147/CLEP.S96797

44. Nacul LC, Lacerda EM, Pheby D, Campion P, Molokhia M, Fayyaz S, Leite JCDC, Poland F, Howe A, Drachler ML. Prevalence of myalgic encephalomyelitis/chronic fatigue syndrome (ME/CFS) in three regions of England: A repeated cross-sectional study in primary care. BMC Med (2011) 9:91. doi:10.1186/1741-7015-9-91

45. Thorley-Lawson DA. EBV Persistence—Introducing the Virus. Curr Top Microbiol Immunol (2015) 390:151. doi:10.1007/978-3-319-22822-8_8

46. Nacul L, O’Boyle S, Palla L, Nacul FE, Mudie K, Kingdon CC, Cliff JM, Clark TG, Dockrell HM, Lacerda EM. How Myalgic Encephalomyelitis/Chronic Fatigue Syndrome (ME/CFS) Progresses: The Natural History of ME/CFS. Front Neurol (2020) 11:826. doi:10.3389/fneur.2020.00826

47. Nacul L, Authier FJ, Scheibenbogen C, Lorusso L, Helland IB, Martin JA, Sirbu CA, Mengshoel AM, Polo O, Behrends U, et al. European Network on Myalgic Encephalomyelitis/Chronic Fatigue Syndrome (EUROMENE): Expert Consensus on the Diagnosis, Service Provision, and Care of People with ME/CFS in Europe. Medicina (Kaunas) (2021) 57:510. doi:10.3390/medicina57050510

48. Sospedra M, Zhao Y, Hausen H Zur, Muraro PA, Hamashin C, De Villiers EM, Pinilla C, Martin R. Recognition of conserved amino acid motifs of common viruses and its role in autoimmunity. PLoS Pathog (2005) 1:0335–0348. doi:10.1371/journal.ppat.0010041

49. Phelan J, Grabowska AD, Sepúlveda N. A potential antigenic mimicry between viral and human proteins linking Myalgic Encephalomyelitis/Chronic Fatigue Syndrome (ME/CFS) with autoimmunity: The case of HPV immunization. Autoimmun Rev (2020) 19:102487. doi:10.1016/j.autrev.2020.102487

50. Rojas M, Restrepo-Jiménez P, Monsalve DM, Pacheco Y, Acosta-Ampudia Y, Ramírez-Santana C, Leung PSC, Ansari AA, Gershwin ME, Anaya JM. Molecular mimicry and autoimmunity. J Autoimmun (2018) 95:100–123. doi:10.1016/j.jaut.2018.10.012

51. Ali S, Matcham F, Irving K, Chalder T. Fatigue and psychosocial variables in autoimmune rheumatic disease and chronic fatigue syndrome: A cross-sectional comparison. J Psychosom Res (2017) 92:1–8. doi:10.1016/j.jpsychores.2016.11.002

52. Moss-Morris R, Chalder T. Illness perceptions and levels of disability in patients with chronic fatigue syndrome and rheumatoid arthritis. J Psychosom Res (2003) 55:305–308. doi:10.1016/S0022-3999(03)00013-8

53. Komaroff AL, Lipkin WI. Insights from myalgic encephalomyelitis/chronic fatigue syndrome may help unravel the pathogenesis of postacute COVID-19 syndrome. Trends Mol Med (2021) 27:895–906. doi:10.1016/j.molmed.2021.06.002

54. Alghamdi M, Alasmari D, Assiri A, Mattar E, Aljaddawi AA, Alattas SG, Redwan EM. An Overview of the Intrinsic Role of Citrullination in Autoimmune Disorders. J Immunol Res (2019) 2019:7592851. doi:10.1155/2019/7592851

55. Birkenfeld P, Haratz N, Klein G, Sulitzeanu D. Cross-reactivity between the EBNA-1 p107 peptide, collagen, and keratin: implications for the pathogenesis of rheumatoid arthritis. Clin Immunol Immunopathol (1990) 54:14–25. doi:10.1016/0090-1229(90)90002-8

56. Pall ML. Levels of Nitric Oxide Synthase Product Citrulline Are Elevated in Sera of Chronic Fatigue Syndrome Patients. J Chronic Fatigue Syndr (2002) 10:37–41. doi:10.1300/J092V10N03_04

57. Naviaux RK, Naviaux JC, Li K, Bright AT, Alaynick WA, Wang L, Baxter A, Nathan N, Anderson W, Gordon E. Metabolic features of chronic fatigue syndrome. Proc Natl Acad Sci U S A (2016) 113:E5472–E5480. doi:10.1073/pnas.1607571113

58. Maes M, Mihaylova I, Leunis J. Chronic fatigue syndrome is accompanied by an IgM-related immune response directed against neoepitopes formed by oxidative or nitrosative damage to lipids. Neuro Endocrinol Lett (2006) 27:615–621. doi:10.1097/YCO.0b013e32831a4728

