## Supplementary Material for "Revisiting IgG antibody reactivity to Epstein-Barr virus in Myalgic Encephalomyelitis/Chronic Fatigue Syndrome and its potential application to disease diagnosis"

### Supplementary Table 1

Brief summary of the EBV proteins analysed and the respective number of 15-mer peptides (antigens) per EBV strain.

| EBV Protein | Associated stage | Number of 15-mer peptides | | | | | | |
|---|---|---|---|---|---|---|---|---|
| | | Overall | AG876 | B95.8 | GD1 | Cao | Raji | P3HR.1 |
| BALF-2 | Early lytic | 290 | 278 | 278 | 278 | 0 | 0 | 0 |
| BALF-5 | Early lytic | 256 | 250 | 250 | 250 | 0 | 0 | 0 |
| BFRF-3 | Late lytic | 42 | 0 | 42 | 0 | 0 | 0 | 0 |
| BLLF-1 | Late lytic | 273 | 204 | 202 | 199 | 0 | 0 | 204 |
| BLLF-3 | Early lytic | 74 | 66 | 67 | 66 | 0 | 0 | 0 |
| BLRF-2 | Late lytic | 41 | 38 | 38 | 38 | 0 | 0 | 0 |
| BMRF-1 | Early lytic | 102 | 99 | 99 | 99 | 0 | 0 | 0 |
| BZLF-1 | Immediate early lytic | 89 | 57 | 57 | 58 | 0 | 0 | 0 |
| EBNA-1 | Latency I, II, and III | 182 | 98 | 107 | 111 | 0 | 0 | 0 |
| EBNA-3 | Latency III | 446 | 223 | 226 | 224 | 0 | 0 | 0 |
| EBNA-4 | Latency III | 469 | 229 | 221 | 224 | 0 | 0 | 0 |
| EBNA-6 | Latency III | 461 | 254 | 234 | 230 | 0 | 0 | 0 |
| LMP-1 | Latency II and III | 197 | 79 | 85 | 80 | 77 | 84 | 0 |
| LMP-2 | Latency II and III | 132 | 120 | 120 | 120 | 0 | 0 | 0 |

**Supplementary Table 2**

Comparison among different null models (included the covariates age and gender and their interaction) using the Akaike's information criterion (AIC). The best model for each analysis (in bold) is the one with the lowest AIC estimate. ME/CFS_all, ME/CFS_inf and ME/CFS_noninf represent all the ME/CFS patients, ME/CFS patients with an infectious trigger, and ME/CFS patients with a non-infectious trigger, respectively

| Analysis/Comparison | Model (Link function) | AIC | ROC (95% CI) |
|---|---|---|---|
| ME/CFS_all vs Healthy Controls | Logit | 189.973 | 0.577 (0.478;0.676) |
| | Probit | 189.964 | 0.576 (0.478;0.675) |
| | **Complementary log-log** | **189.936** | **0.574 (0.475;0.672)** |
| ME/CFS_inf vs Healthy Controls | Logit | 147.055 | 0.610 (0.500;0.719) |
| | **Probit** | **147.029** | **0.606 (0.496;0.715)** |
| | Complementary log-log | 147.220 | 0.609 (0.499;0.718) |
| ME/CFS_noninf vs Healthy Controls | Logit | 127.619 | 0.556 (0.429;0.683) |
| | Probit | 127.629 | 0.559 (0.432;0.687) |
| | **Complementary log-log** | **127.547** | **0.556 (0.429;0.683)** |
| ME/CFS_inf vs ME/CFS_noninf | **Logit** | **129.205** | **0.596 (0.471;0.720)** |
| | Probit | 129.236 | 0.597 (0.472;0.721) |
| | Complementary log-log | 129.529 | 0.596 (0.472;0.721) |

**Supplementary Table 3**

The top 5 most significant antibodies for each association analysis where the adjusted p-value is shown within brackets where ME/CFS_all, ME/CFS_inf and ME/CFS_noninf represent all ME/CFS patients, ME/CFS patients with an infectious trigger, and ME/CFS patients with a non-infectious trigger, respectively. For simplicity, the antibodies w

ere labelled according to their peptide. Statistically significant findings were obtained for $-\log_{10}$(adjusted p-value) > 1.30 (=$-\log_{10}(0.05)$) controlling for false discovery rate of 5%.

| **Analysis** | **Peptide** | **$-\log_{10}$(adjusted p-value)** |
|---|---|---|
| ME/CFS_all vs Healthy controls | EBNA6_0066 | 0.743 |
| | BLRF2_0005 | 0.486 |
| | EBNA4_0392 | 0.486 |
| | EBNA4_0497 | 0.486 |
| | EBNA4_0529 | 0.486 |
| ME/CFS_inf vs Healthy controls | EBNA6_0066 | 2.693 |
| | EBNA6_0070 | 2.693 |
| | EBNA4_0529 | 1.794 |
| | EBNA3_0380 | 1.270 |
| | EBNA6_0569 | 1.270 |
| ME/CFS_noninf vs Healthy controls | EBNA6_0782 | 1.193 |
| | BALF2_0358 | 1.153 |
| | BALF2_0765 | 1.153 |
| | BALF5_0041 | 1.153 |
| | BALF5_0206 | 1.153 |

**Supplementary Figure 1**

Distributions of the Spearman's correlation coefficient between all the possible pairs of EBV-derived antibodies in healthy controls, all ME/CFS patients, ME/CFS patients with an infectious trigger, and ME/CFS patients with a non-infectious or unknown trigger.

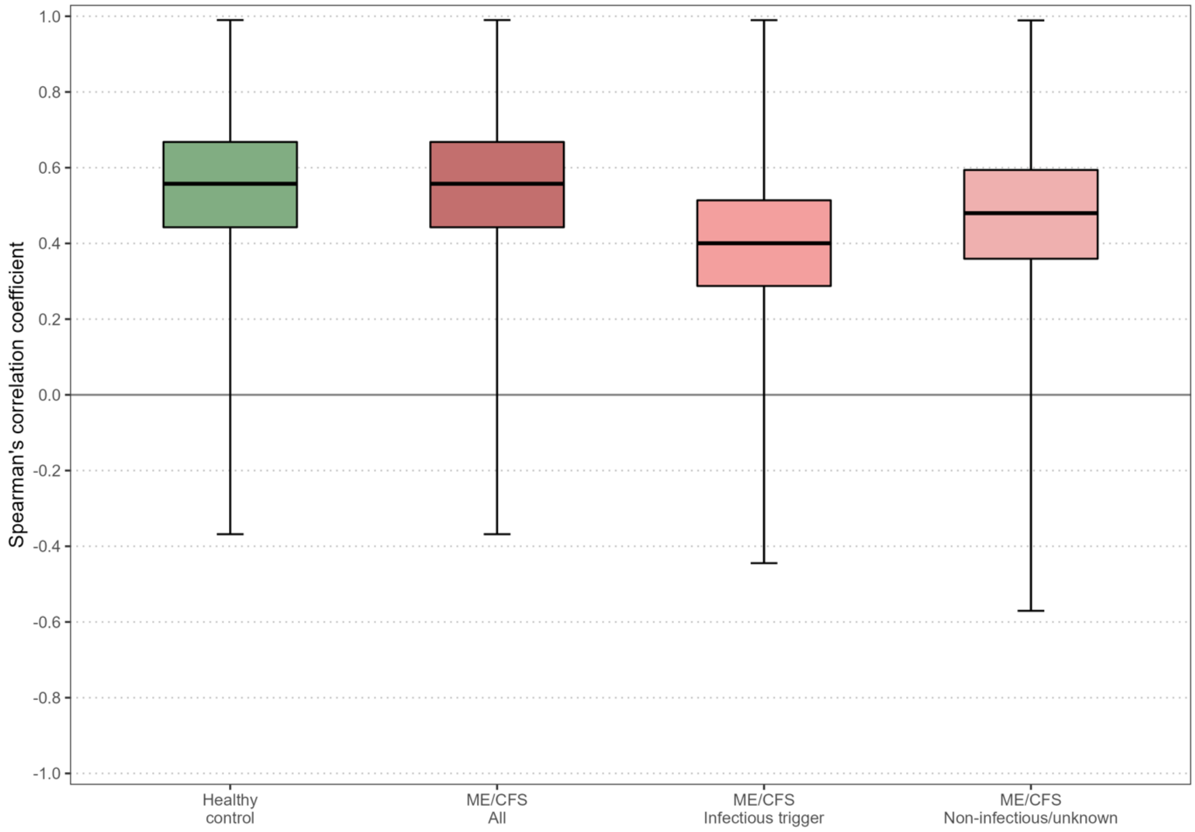
